# Evaluation of the costs of care for pediatric patients hospitalized for RSV: a retrospective cohort study in Belgium

**DOI:** 10.1101/2024.11.21.24317618

**Authors:** Anne Tilmanne, Magali Pirson, Pol Leclercq, Julie Van Den Bulcke, Arnaud Bruyneel

## Abstract

**Objectives:** This study aimed to evaluate the costs of respiratory-syncytial virus (RSV)-related hospitalizations in children under 3 years old in Belgium for hospitals and health insurance, and to identify factors influencing costs.

**Study design:** This retrospective cohort study used data from 16 French-speaking hospitals in Belgium, covering January 1, 2018, to December 31, 2019. RSV diagnoses for children under three were identified using International Classification of Diseases-Tenth Revision (ICD-10) codes, resulting in 2,176 hospitalizations analyzed for cost assessment. Hospital and health insurance costs were derived from administrative and billing data, adjusted for inflation, and analyzed using descriptive and inferential statistics, including regression models to assess cost factors.

**Results:** RSV prevalence among pediatric hospitalizations peaked at 12.9% in 2018 and 10.0% in 2019, with children under 3 years showing a prevalence of 16.7% both years. Of the 2,176 RSV-related hospitalizations, 61.8% were in children under one year, and 28.6% had readmissions within a year. The median length of stay (LOS) was 3.67 days, with a median hospital cost of €2,924 and a median health insurance cost of €2,221 per stay. Factors associated with higher costs included longer LOS, severe diagnosis-related group category, pediatric intensive care unit admission, and non-invasive ventilation use, with costs generally lower for children aged 1-2 years.

**Conclusions:** This study highlights the significant financial burden of RSV hospitalization in Belgium, with costs exceeding €25 million annually. This emphasizes the need for preventive measures and better resource allocation to reduce the economic impact of RSV on healthcare systems.

## Introduction

Respiratory syncytial virus (RSV) is one of the most feared viral infections in infants, and almost all children will be infected by 2 years of age.^1^ A recent European, multicenter, prospective, observational birth cohort study in healthy term-born infants revealed an incidence of RSV infection of 26.2% and a rate of medically-attended RSV infection of 14.1%, during the first year of life.^2^ While over 97% of deaths attributed to RSV occur in low- and middle-income countries, the healthcare impact of RSV infection is significant in high-income nations as well, where it varies according to age and by country. In the USA, the estimated annual hospitalization rate is three per 1000 children under 5 years old.^3^ In Europe, a study based on International Classification of Diseases-Tenth Revision (ICD-10) coded hospitalizations showed that the average number of hospitalizations for RSV ranges from 20.5 to 22.3 per 1000 children under one year of age in Scotland, Finland, and Denmark, and ranges from 8.6 to 11.7 per 1000 children in England, the Netherlands, and Italy.^4^ In Belgium, hospitalization rates are higher with 68.3 hospitalizations per 1000 children under 1 year of age and 5.0 per 1000 children aged 1–4 years in 2018.^5^ Intensive care unit (ICU) admission is required for 5% of the hospitalized children in Europe.^2,5^ Compared with the average in children under 5 years, newborns have a higher risk of being admitted to the ICU (15.9% vs. 5.1%).^5^

There is no treatment available against RSV. Until 2023, the only possibility against RSV in Belgium was palivizumab, a monoclonal antibody active against RSV but in prophylaxis only. The cost of a vial is 376 euros, and monthly injections are required during the RSV season, given the molecule’s short half-life. This passive immunization is reimbursed by social security to an at-risk population in Belgium: children born at 28 weeks of gestation or less who are less than 1 year old at the beginning of the season; children born between 28 and 35 weeks of gestational age who needed more than 48 hours of ventilation, stayed in a neonatal intensive care unit (NICU) center, and are less than 6 months of age at the onset of the RSV season; and children less than 2 years of age with hemodynamically significant congenital heart disease or who require chronic oxygen supplementation/ventilation.^6^

Since January 2024, a maternal RSV vaccine has been available in Belgium to prevent lower respiratory tract disease. The vaccine has a reported efficacy rate of 69.4% (97.58% confidence interval [CI], 44.3 to 84.1) within 180 days after birth for preventing medically-attended severe RSV-associated lower respiratory tract illness and 56.8% (99.17% CI, 10.1 to 80.7) for preventing RSV-associated hospitalization.^7^ As of November 1, 2024, nirsevimab—a monoclonal antibody that targets the RSV fusion protein with an extended half-life—is available in Belgium for the passive immunization of newborns. One dose is reimbursed by social security for infants under 6 months of age. The efficacy of nirsevimab in children 12 months of age or younger for preventing hospitalization due to RSV-associated lower respiratory tract infection is 83.2% (95% CI: 67.8 to 92.0), and 75.7% (95% CI: 32.8 to 92.9) for preventing very severe RSV-associated lower respiratory tract infection.^8^

Continuous surveillance data are lacking to quantify the impact of RSV on children and their families in Belgium (e.g., severity of illness, need for hospitalization, need for oxygen supplementation, absence from work), as well as on healthcare systems (e.g., medical consultations, length of hospital stay, ICU admissions, type and duration of respiratory support).^9^ However, these elements can be found in the minimum clinical data set (MCD) required for each hospitalization.^10^ Additionally, data on the costs of patients hospitalized with RSV are missing, and an assessment of the economic burden of RSV is rarely available at the national level.^11^ The absence of comprehensive data hinders the ability to develop targeted public health strategies and allocate resources efficiently. This information can then be used to evaluate the cost-effectiveness of prevention programs, such as vaccination campaigns, and new treatments available on the market.^12–15^ Improved data collection and analysis would provide a clearer picture of the burden of RSV, enabling better healthcare planning and policymaking to mitigate its effects on public health.^10^

The primary objective of this study was to evaluate costs for hospitals and health insurance of RSV-associated hospitalizations in children under 3 years of age in Belgium. The secondary objective was to identify factors that influence those costs.

## Materials and methods

### Study design and setting

This retrospective cohort study used data from 16 French-speaking hospitals in Belgium, covering the period from 1 January, 2018 to 31 December, 2019. These 16 hospitals accounted for approximately 10% of all pediatric hospitalizations in Belgium and 30% of pediatric hospitalizations in the French-speaking part of the country for both years. The Strengthening Reporting of Observational Studies in Epidemiology (STROBE) guidelines for cohort studies were applied to this study.^16^

### Participants

To calculate prevalence by month, all pediatric unit admissions (without NICU) under the age of 3 years (n=93,604) and 16 (n=97,821) were considered across the 16 hospitals. The diagnosis of RSV was determined based on medical discharge summaries, coded with ICD-10 codes J205, J210, and J121 for the primary diagnosis and code B974 for secondary diagnoses. A total of 243 records with incomplete stays (e.g., no administrative data found in the hospital, very short stay <6 hours, no data on costs per pathology, and still in the hospital on December 31, 2019) and 75 patients who were older than 3 years of age were excluded. In total, data for 2,176 hospitalizations for patients with a diagnosis of RSV were analyzed in the study.

### Variables and data sources

The data set included information on inpatients that was extracted from medical discharge summaries, administrative data (e.g., length of stay (LOS), admission to pediatric intensive care unit [PICU]) and medical billing and accountancy files. Two types of costs were evaluated: those from the perspective of the hospital and those from the health insurance perspective, with an analysis per stay for each perspective.

The costs from the hospital perspective were determined using a full cost approach with an analytical cost accounting methodology.^17,18^ The full costing approach included physicians costs, nursing costs, drug costs, consumable costs, equipment costs, radiology costs, and laboratory costs. Each expense was allocated per patient using a specific key (e.g., the cost of nursing was allocated based on the LOS and the time spent on care). The health insurance perspective was derived from hospital billing files and reflected costs to be paid by the health insurance fund to hospitals. These costs included: 1) medical procedures, including laboratory tests and medical imaging, 2) drug costs, and 3) lump sums and other miscellaneous expenses. Additionally, the Budget of Financial Means (BFM), which covers administrative costs, hotel costs, investments, nursing care, and other related expenses, was included. All costs, from both perspectives, were calculated individually and for the entire duration of the stay. The 2018 costs were adjusted for inflation in Belgium in 2019 (2.05%), and all costs in the study are expressed in euros (€).

Some of the variables were taken from the ICD-10 classification, which is an internationally used list of diseases defined by the World Health Organization. All Belgian hospitals are required to communicate this data two times per year as part of their funding. From these ICD-10 codes, the diagnosis-related groups (DRGs) were obtained via the 3M grouper, which uses the All Patients Refined DRG (APR-DRG) system that includes 355 different classes.^19^ The Severity of Illness (SOI) is determined based on the DRG complications hierarchy, which classifies the extent of organ system loss-of-function or physiologic decompensation on a scale of minor (1), moderate (2), major (3), or extreme (4). Patient assignment to these subgroups considers not only specific secondary diagnoses but also interactions between secondary diagnoses, age, principal diagnosis, and procedures.^20^ SOI determination is disease-specific, with high severity of illness primarily resulting from interactions between multiple conditions. The risk of mortality indicates the likelihood of death. A patient may exhibit major severity of illness but only a minor risk of mortality.

Sociodemographic data (e.g., age, sex, mortality) were extracted from the minimum hospital discharge summary and administrative data.^21^ Data on medical procedures (e.g., monitoring, non-invasive ventilation, peripheral infusion, nasogastric tube, supplementary oxygen) were sourced from invoicing files.

### Statistical methods

Statistical analyses were conducted using STATA® version 18 and R software version 4.1.2 (R Core Team). A p-value < 0.05 was considered statistically significant. Descriptive statistics are presented as proportions for categorical variables, means with standard deviations (SD) for continuous variables, and medians with interquartile range (IQR) for non-normally distributed variables. Mann-Whitney tests were used for asymmetric variables, while Student’s T tests and chi-square tests were employed for normally distributed variables and proportions. The Kolmogorov-Smirnov test assessed the symmetry of continuous variables.

Linear regression models were performed on log-transformed hospital and health insurance costs, to normalize the distribution of residuals. Additionally, the LOS was categorized into quartiles. When the assumption of homoscedasticity was not met, robust standard errors were computed to control for heteroscedasticity. In the univariate analysis, the raw geometric means of hospital costs and the coefficients from linear regression were computed for each indicator. The multivariate model takes into account all the factors available in the database. To appreciate the adjusted effect of each predictor on the dependent variable in the multivariate model, we exponentiated the coefficients to obtain the adjusted ratio of geometric mean (GMR). For example, a specific category with a GMR of 1.50 was interpreted as a hospital cost 50% greater compared to the reference group.

Univariate logistic regression models were used to explore associations between independent variables (i.e., socio-demographic data, data from minimum hospital discharge summaries, administrative data, and invoicing data) and dependent variables (i.e., cost of hospital and health insurance). Variables with a p-value < 0.05 in univariate analyses were included in multiple logistic regression models.

### Ethical considerations

The inpatient records used in the retrospective study were fully anonymized by the hospitals and the research team did not have any access to medical files. The study was approved by the Ethics Committee of an academic hospital in Belgium (reference: N2023/047).

## Results

### Prevalence of RSV among hospitalized children

In 2018, the peak prevalence of RSV infection among pediatric hospitalizations reached a maximum of 12.9% overall and 16.7% for children under three years of age. In comparison, the figures for 2019 were 10.0% and 16.7%, respectively. An increase was observed from October to March, with a peak in December for both years (Figure 1).

**Figure 1.**
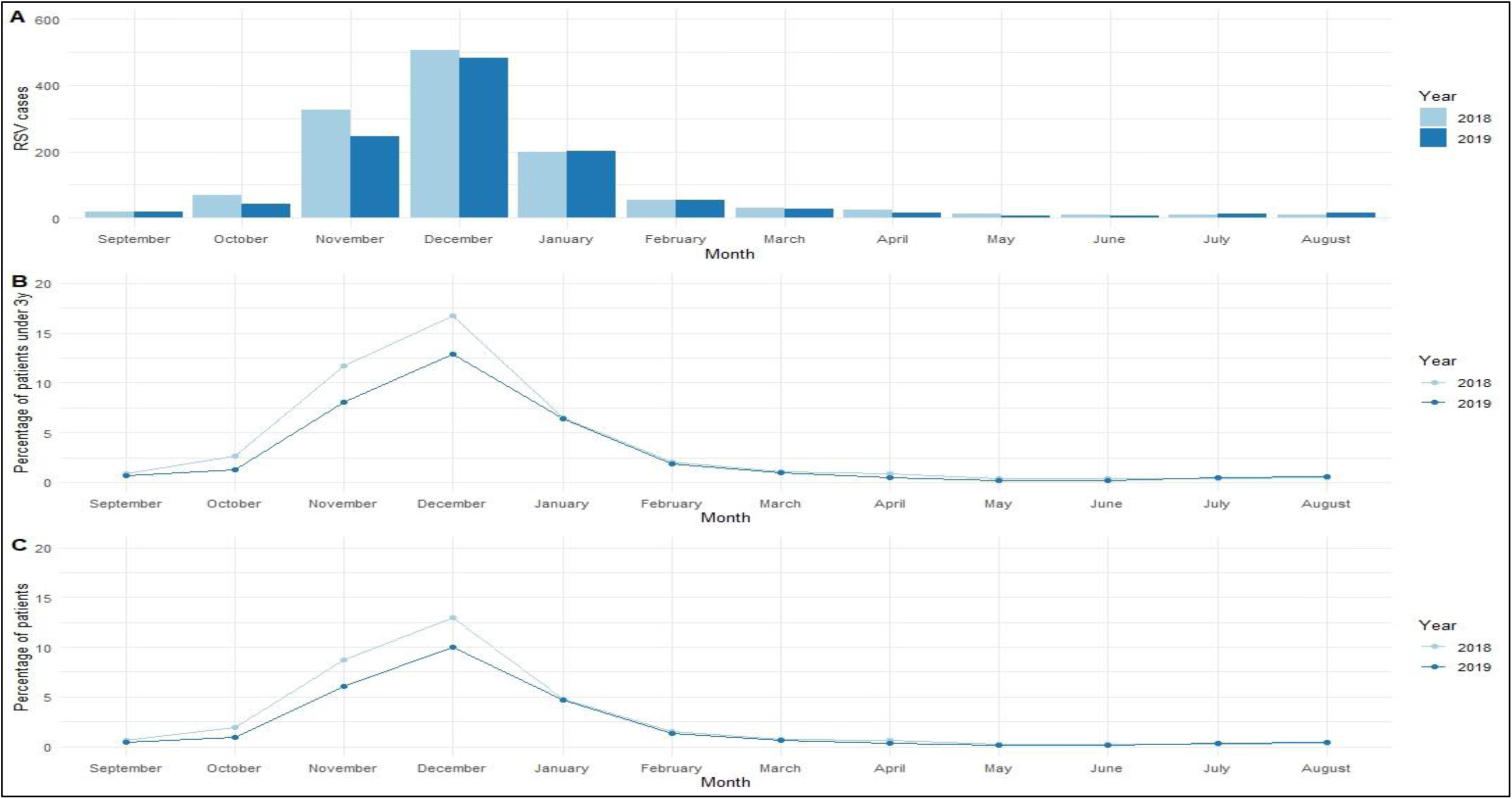
Monthly and yearly number of hospitalizations due to RSV infection in the cohort (A), percentage of RSV cases among hospitalized children under 3 years old (B) and those under 16 years old (C).

### Patient characteristics

During the two-year period covered by the available data, a total of 2,176 hospitalizations were reported among children under three years old, with RSV identified as either the primary (1921 cases) or secondary (255 cases) reason for hospitalization. Children under one year old comprised the majority of the cohort, accounting for 1,345 out of 2,176 hospitalizations (61.8%), followed by children aged one to two years old with 703 hospitalizations (32.3%), and children aged two to three years old with 128 hospitalizations (5.9%). The median [p25-p75] LOS was 3.67 [2.2-5.4] days. Ninety-six patients (4.4%) required admission to a pediatric ICU, 110 patients (5.1%) needed non-invasive ventilation, and 178 patients (8.2%) experienced hypoxemia during hospitalization. There was one reported death. Readmission to the same hospital within the year was observed in 623 cases (28.6%). Between the two years, a significantly higher percentage of patients in the 0 to 1 year age category was observed in 2018 (64.8% vs 58.4%, p-value: 0.006). Overall, the clinical severity of the patients was also significantly lower in 2018, with fewer patients in the major APR-DRG severity category (19.2% vs 23.9%, p-value: 0.003), less hypoxemia (7.2% vs 9.3%, p-value: 0.042), less use of monitoring (82.8% vs 89.3%, p-value: <0.0001), fewer peripheral perfusions (41.4% vs 45.9%, p-value: 0.033), and fewer nasogastric tube use (17.0% vs 24.2%, p-value: <0.0001) (Table 1).

**Table 1.** Patient Characteristics.

| Characteristics | Overall<br>(n=2,176) | 2018<br>(n=1,155) | 2019<br>(n=1,021) | pvalue |
| --- | --- | --- | --- | --- |
| **Inpatient RSV coding, n (%)** |  |  |  |  |
| *Primary diagnosis* | 1,921 (88.3) | 1,023 (88.6) | 838 (82.1) | 0.654 |
| *Secondary diagnosis* | 255 (11.7) | 132 (11.4) | 123 (12.0) |  |
| **Age category, n (%)** |  |  |  |  |
| *0-1* | 1,345 (61.8) | 749 (64.8) | 596 (58.4) | 0.006 |
| *1-2* | 703 (32.3) | 347 (30.0) | 356 (34.9) |  |
| *2-3* | 128 (5.9) | 59 (5.1) | 69 (6.8) |  |
| **Gender, n (%)** |  |  |  |  |
| *Girl* | 996 (45.8) | 537 (46.5) | 459 (45.0) | 0.472 |
| *Boy* | 1,180 (54.2) | 618 (53.5) | 562 (55.0) |  |
| **Emergency department admission, n (%)** | 1,960 (90.1) | 1,036 (89.7) | 924 (90.5) | 0.532 |
| **LOS (days), median [p25-p75]** | 3.7<br>[2.2-5.4] | 3.6<br>[2.2-5.1] | 3.7<br>[2.2-5.6] | 0.242 |
| **APR-DRG, n (%)** |  |  |  |  |
| *138 - Bronchiolitis and respiratory syncytial virus (RSV) pneumonia* | 1,517 (69.7) | 791 (68.5) | 721 (70.6) | 0.205 |
| *113 - Upper respiratory tract infections* | 174 (8.0) | 84 (7.3) | 90 (8.8) |  |
| *144 - Respiratory signs, symptoms, and minor diagnoses* | 115 (5.3) | 61 (5.3) | 54 (5.3) |  |
| *951 - Moderately extensive procedure unrelated to the main diagnosis* | 185 (8.5) | 106 (9.2) | 79 (7.7) |  |
| *Others* | 185 (8.5) | 111 (9.6) | 77 (7.5) |  |
| **APR-DRG severity category, n (%)** |  |  |  |  |
| *Minor* | 856 (39.3) | 489 (42.3) | 367 (35.9) | 0.003 |
| *Moderate* | 822 (37.8) | 423 (36.6) | 399 (39.1) |  |
| *Major* | 466 (21.4) | 222 (19.2) | 244 (23.9) |  |
| *Extreme* | 32 (1.5) | 21 (1.8) | 11 (1.1) |  |
| **APR-DRG mortality risk category, n (%)** |  |  |  |  |
| *Low/Moderate* | 2,156 (99.1) | 1,144 (99.0) | 1,012 (99.1) | **0.014** |
| *High/Extreme* | 20 (0.9) | 11 (1.0) | 9 (0.9) |  |
| **PICU admission, n (%)** | 96 (4.4) | 55 (4.8) | 41 (4.0) | **0.004** |
| **Mortality, n (%)** | 1 (0.1) | - | - | - |
| **Transfer to another hospital, n (%)** | 83 (3.8) | 40 (3.5) | 43 (4.2) | 0.363 |
| **Readmission, n (%)** | 1,153 (53.0) | 832 (72.0) | 721 (70.6) | 0.465 |
| **Hypoxemia, n (%)** | 178 (8.2) | 83 (7.2) | 95 (9.3) | **0.042** |
| **Non-invasive ventilation, n (%)** | 110 (5.1) | 59 (5.1) | 52 (5.1) | 0.940 |
| **Monitoring, n (%)** | 1,868 (85.8) | 956 (82.8) | 912 (89.3) | **<0.0001** |
| **Peripheral infusion, n (%)** | 947 (43.5) | 478 (41.4) | 469 (45.9) | **0.033** |
| **Nasogastric tube, n (%)** | 443 (20.4) | 196 (17.0) | 247 (24.2) | **<0.0001** |

### Cost of RSV

The median cost per RSV-related hospital stay borne by the hospital was €2,924 [1,919-4,404], while the median cost borne by health insurance was €2,221 [1,796-2,852] (Figure 2).

**Figure 2.**
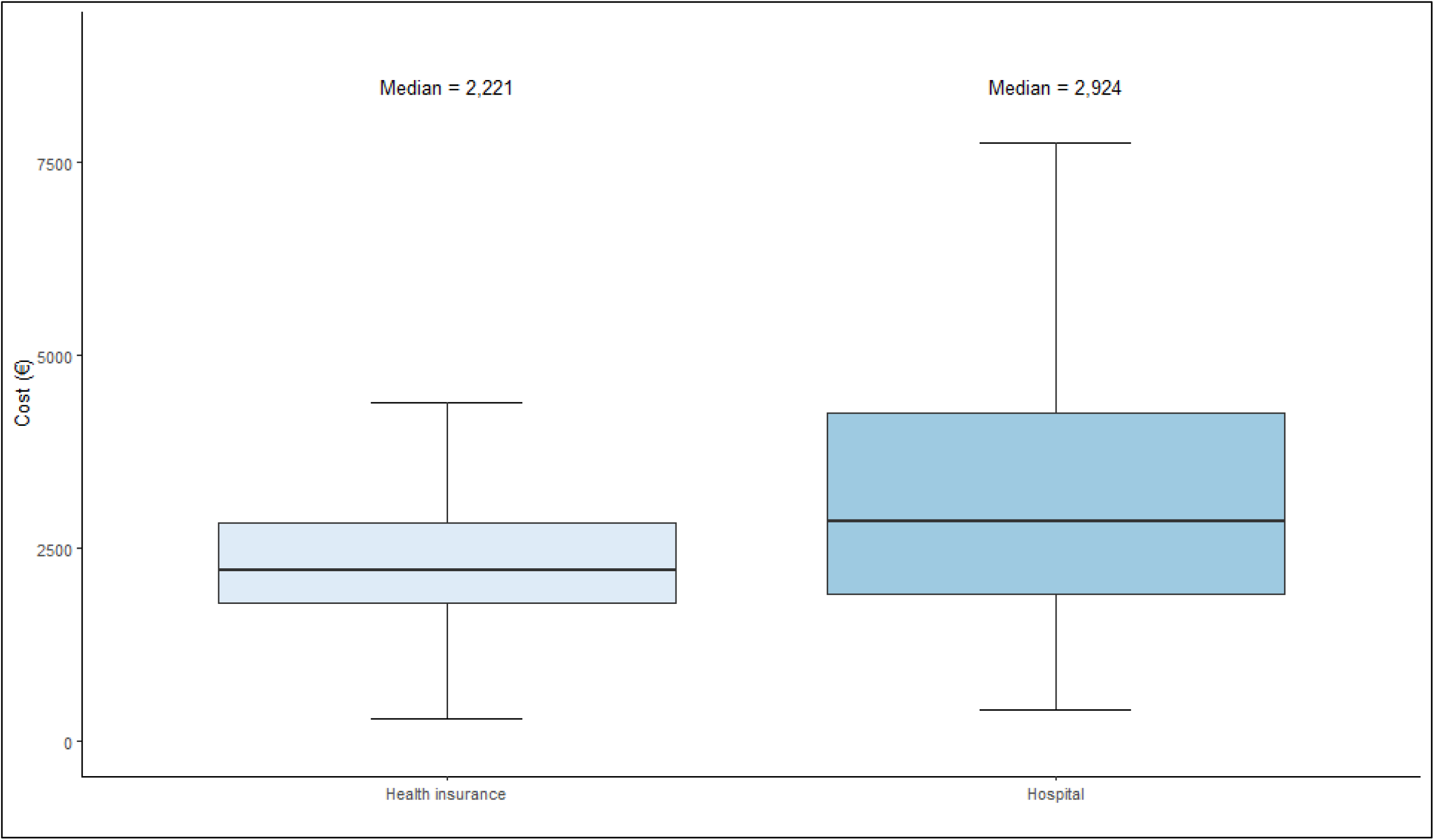
Box plot comparing the cost to the hospital and cost to health insurance of patients hospitalized for RSV.

### Factors associated with RSV costs

For univariate analyses, from the hospital perspective, the median costs were higher for patients with LOS exceeding 4 days, at €5,790 [€4,625-€7,208], extreme APR-DRG category at €7,295 [€5,340-€10,181], admission to PICU at €7,295 [€5,340-€10,181], and those receiving non-invasive ventilation at €5,602 [€3,637-€7,968]. From the health insurance perspective, the trends were similar to the hospital perspective with respective median costs of €3,267 [€2,889-€4,019], €4,050 [€2,935-€7,656], €4,364 [€3,740-€5,725], and €3,984 [€3,027-€4,851] (Appendix Table 1).

For multivariable factors associated with hospital costs, in 2019 compared to 2018, longer LOS, higher severity categories of APR-DRG, admission to PICU, use of non-invasive ventilation, and placement of peripheral infusion were significantly associated with higher costs. Similarly, for health insurance costs, longer LOS, extreme severity category of stay, admission to PICU, and placement of infusion were significantly associated with higher costs. Conversely, patients aged 1-2 years compared to those under 1 year had lower costs (Figure 3).

**Figure 3.**
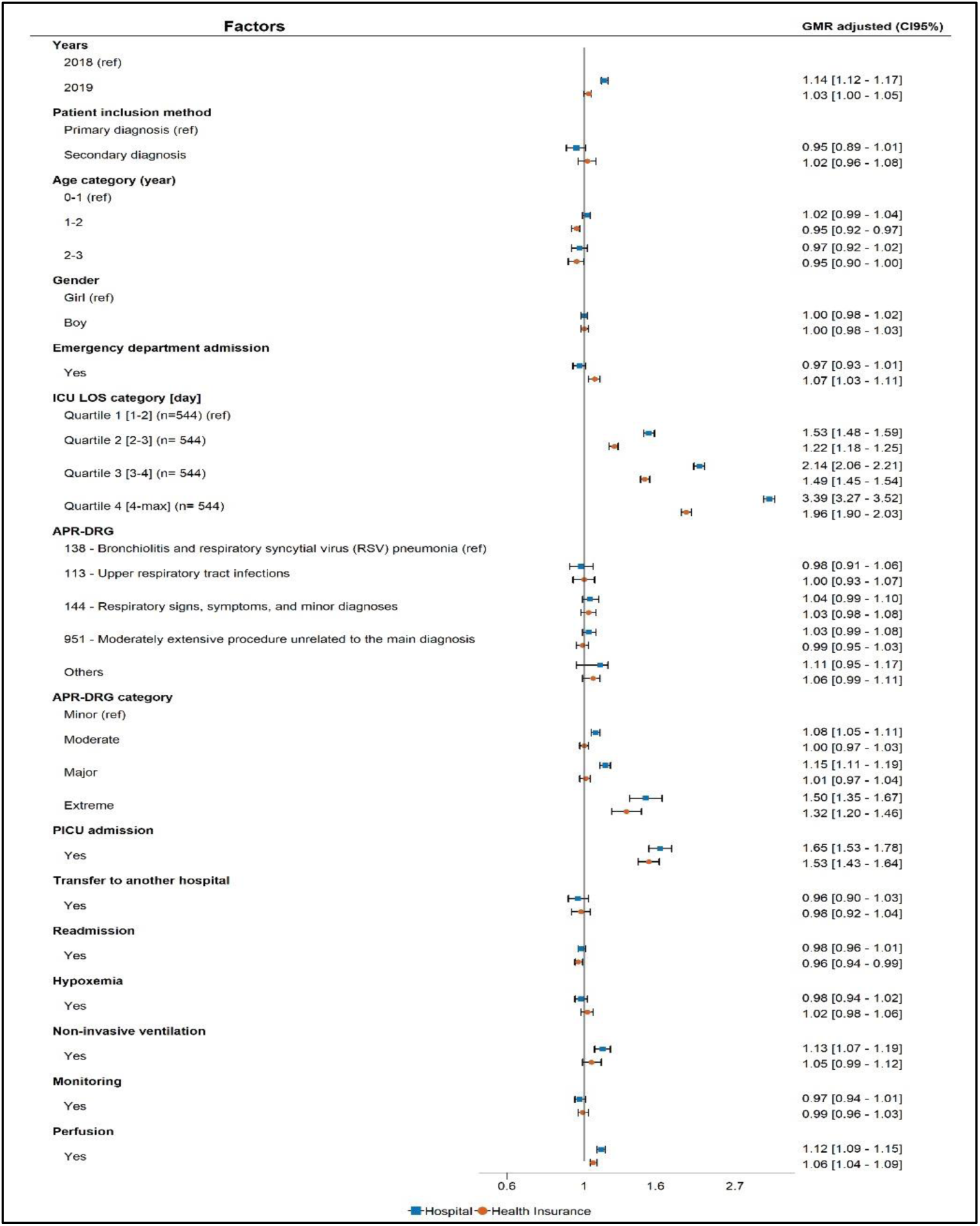
Factors associated with hospital and social health insurance costs in the multivariable model.

## Discussion

To our knowledge, this is the first study to analyze hospitalization costs in Belgium for children under 3 years old with RSV. The particular strength of this work lies in its dual analysis approach. The first analysis involved evaluating the costs associated with RSV hospitalization from data extracted from social security records (“health insurance” in the tables). The second analysis was based on data directly obtained from hospitals (“hospital” in the tables), allowing for a more accurate and realistic analysis, as well as a comparison between the two sources.

Although the present work represents only a proportion of RSV hospitalizations in Belgium, the findings are consistent with the Belgian 2018 national data reported by Bouckaert et al.^5^ In the present study, among the participating hospitals, RSV hospitalizations in 2018-2019 accounted for 3.5% of the annual pediatric ward admissions, rising to 16.7% during the peak RSV activity in November-December for children under three years of age. Bouckaert et al.^5^ demonstrated an annual pediatric bed occupancy rate of 5% due to RSV, rising to as high as 30% during the seasonal peak in November-December. Bouckaert et al. also reported five RSV-linked in-hospital deaths (5/10,545 hospitalizations, or 0.46 deaths per 1000 hospitalizations), which aligns with the findings of the present study (1/2176, or 0.46 deaths per 1000 hospitalizations). PICU admission rates, the proportion of major and extreme SOI cases and the median LOS are comparable between the two studies.^5^

Regarding the analysis of costs per stay covered by health insurance, the median cost per stay was €2,221 in this study. This is comparable to other European studies, although variations— both minor and significant—can be observed. In Germany, Niekler et al.^22^ reported median costs of €2,389 for children under 1 year and €2,690 for children aged 1 to 4 years. In France, Demont et al.^23^ calculated a mean cost with significant variation based on age: from €1,448 for children aged 24-35 months to €2,953 for those under 3 months. In another French study in Lyon,^11^ the mean cost of RSV hospitalization was calculated at €3,973, which is €1,000-2,000 higher than the study from Demont et al.^23^ in the same country. In the USA, higher costs were reported, with mean costs for hospitalization in the first year ranging from $8,324 to $89,000.^24^

Variations in cost between studies can be attributed to differences in study methodologies. In the present work, the calculated median cost per hospitalization was €2,221, based on expenses covered by health insurance, but increased to €2,924 when calculated using actual hospital costs. The likely underestimation of hospitalization costs in studies based solely on reported costs reimbursed by health insurance has been acknowledged by others, such as Niekler et al. in Germany^22^ and Demont et al. in France.^23^ This type of cost calculation often fails to cover the actual expenses incurred by hospitals, as demonstrated in the present study. Expenses included in healthcare costs can vary significantly depending on the country and the insurance policies in place. In Belgium, pediatric healthcare is known for being underfunded and is considered not very profitable in a financial point of view. Providing care for a child requires reassurance and the presence of multiple people; even for a simple blood draw, at least two additional staff members beyond the parents can be necessary, and sometimes three. What might appear to be a quick, simple procedure can often take considerable time. But time-consuming tasks such as feeding, comforting anxious parents, and providing extra surveillance are not fully accounted for by health insurance. Furthermore, certain expenses, like cardio-respiratory monitoring beyond 48 hours or supplemental oxygen provided outside of a PICU setting, are not billable in pediatric wards.

Patient risk factors also significantly impact hospital costs, as highlighted by McLaurin et al.^24^ in the USA. That study reported that the mean first-year hospitalization costs for RSV ranged from $8,324 to $89,000, with higher costs associated with pre-term infants or those with comorbidities. A discussion about other potential risk factors that could affect costs—such as prematurity, age in months rather than years, and the presence of comorbidities—was not possible in this analysis, as these specific data were, unfortunately, not available in the Belgian database.

Logically, the severity of the disease influences the median costs reported by hospitals and health insurance in the present study. This is reflected by LOS, APR-DRG severity, and PICU admission. Niekler et al.^22^ reported similar findings, though a direct comparison of the cost increase is challenging because their study included both pediatric and adult patients.

Costs can also fluctuate based on the year and geographic location. The hospitalization data for RSV analyzed for the years 2018-2019 in the 16 participating hospitals shows a significant difference in age group distribution in 2019 compared to 2018. There were fewer cases in 2019 (1,021) compared to 2018 (1,155), mainly due to a decrease in hospitalizations among children under 1 year old. This variation in the severity, timing, and duration of RSV seasons is well-documented worldwide, though the underlying causes are not fully understood. Factors such as viral genotype, virulence factors, risk factor distribution, altitude, and weather may play a role, but studies attempting to explain these variations have produced conflicting results.^27–30^ These temporal and geographical variations may account, in part, for differences in the median cost of RSV hospitalization.

Based on our cost calculations and the incidence data from the Belgian national study,^5^ it is possible to estimate the annual financial burden of hospitalizations due to RSV for the national health insurance in Belgium. According to Bouckaert et al.,^5^ in 2018, 10,545 hospital episodes were recorded in children under 5 years old, with 8,046 of those involving infants under 1 year old. Based on our average cost estimate for RSV-related hospitalizations (€2,450 ± €1,604), this amounts to approximately €25,835,250, with a variation ranging from €8,921,070 to €42,749,430. Of this amount, 76.4% (€19,712,700 ± €12,905,784) is due to hospitalization of infants under 1 year old. Although these are approximate figures which do not account for inflation in recent years, they provide a strong indication of the significant financial strain RSV places on healthcare systems. Furthermore, this estimate does not include potential long-term costs, such as those associated with chronic respiratory conditions following severe RSV infections and indirect costs, which could further increase the overall economic burden. These findings highlight the importance of healthcare planning and resource allocation to better manage the impact of RSV on both the healthcare system and public health.

## Limitations

This study has several limitations. First, the variables used were collected retrospectively from hospital databases, rather than prospectively. Patient-related variables are derived from billing data, which may not always accurately reflect the actual care provided. Second, cost comparisons should be interpreted with caution, as differing methodologies and perspectives may influence the calculations. Third, the use of anonymized administrative data prevents the collection of information on patient medical severity, which is both a risk factor and a cost predictor for hospitals. Additionally, the quality of diagnostic coding in hospitals presents another limitation. Fourth, the data is somewhat dated, requiring adjustments for inflation and changes in medical practices. Lastly, care should be taken in interpreting the results, as hospital accounting data may contain errors.

## Conclusion

The present study provides valuable insights into the hospitalization costs associated with RSV in children under 3 years old in Belgium, marking the first comprehensive analysis of its kind. By analyzing costs based on social security records and actual hospital data, the present study highlights significant discrepancies between these sources, with hospital-based calculations revealing higher, more accurate cost estimates. This discrepancy underscores the limitations of relying solely on health insurance data for evaluating healthcare expenses. The median cost of an RSV-related hospital stay was €2,221 based on health insurance data and €2,924 when calculated using hospital-reported costs.

The financial burden of RSV hospitalization is substantial, with costs estimated to exceed €25 million annually for Belgian national health insurance. These findings underline the importance of implementing preventive measures, such as vaccination, and ensuring appropriate resource allocation to adequately fund these preventive measures as well as the pediatric departments that care for RSV infected patients. Future research should focus on long-term costs, such as chronic respiratory conditions and indirect economic burdens, to provide a more complete picture of RSV’s financial impact.

## Data Availability

The dataset used and analyzed in this manuscript is available from the corresponding author upon reasonable request.

## Acknowledgments

The authors acknowledge the contribution of a medical writer, Sandy Field, PhD, to the preparation of this manuscript.

## Abbreviations

APR-DRG: all patients refined disease-related group

CI: confidence interval

DRG: disease-related group

ICU: intensive care unit;

ICD-10: International Classification of Diseases-10^th^ revision

IQR: interquartile range

LOS: length of stay

NICU: neonatal intensive care unit

PICU: pediatric intensive care unit

RSV: respiratory syncytial virus

SOI: severity of illness

---

**Appendix Table 1.**
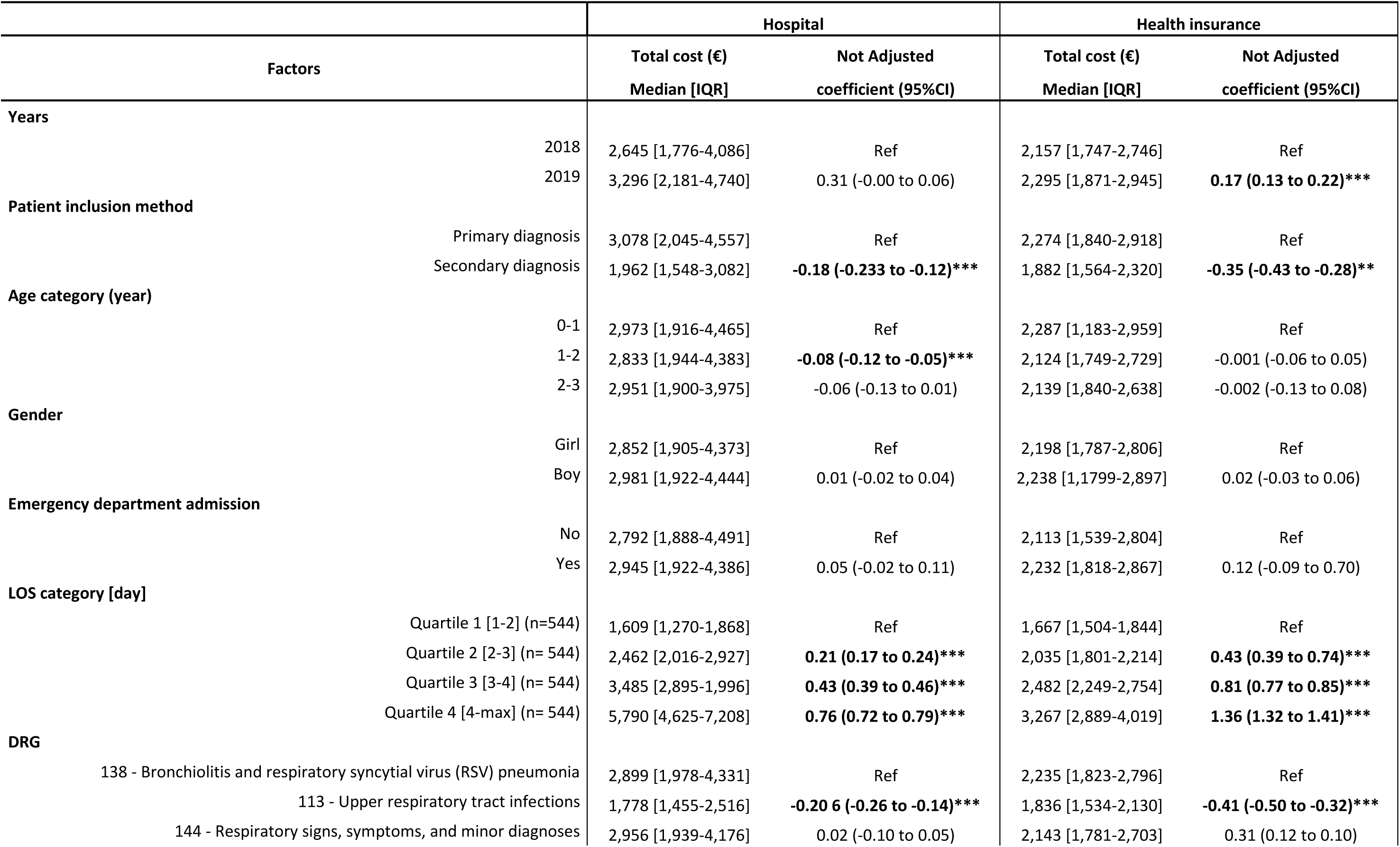
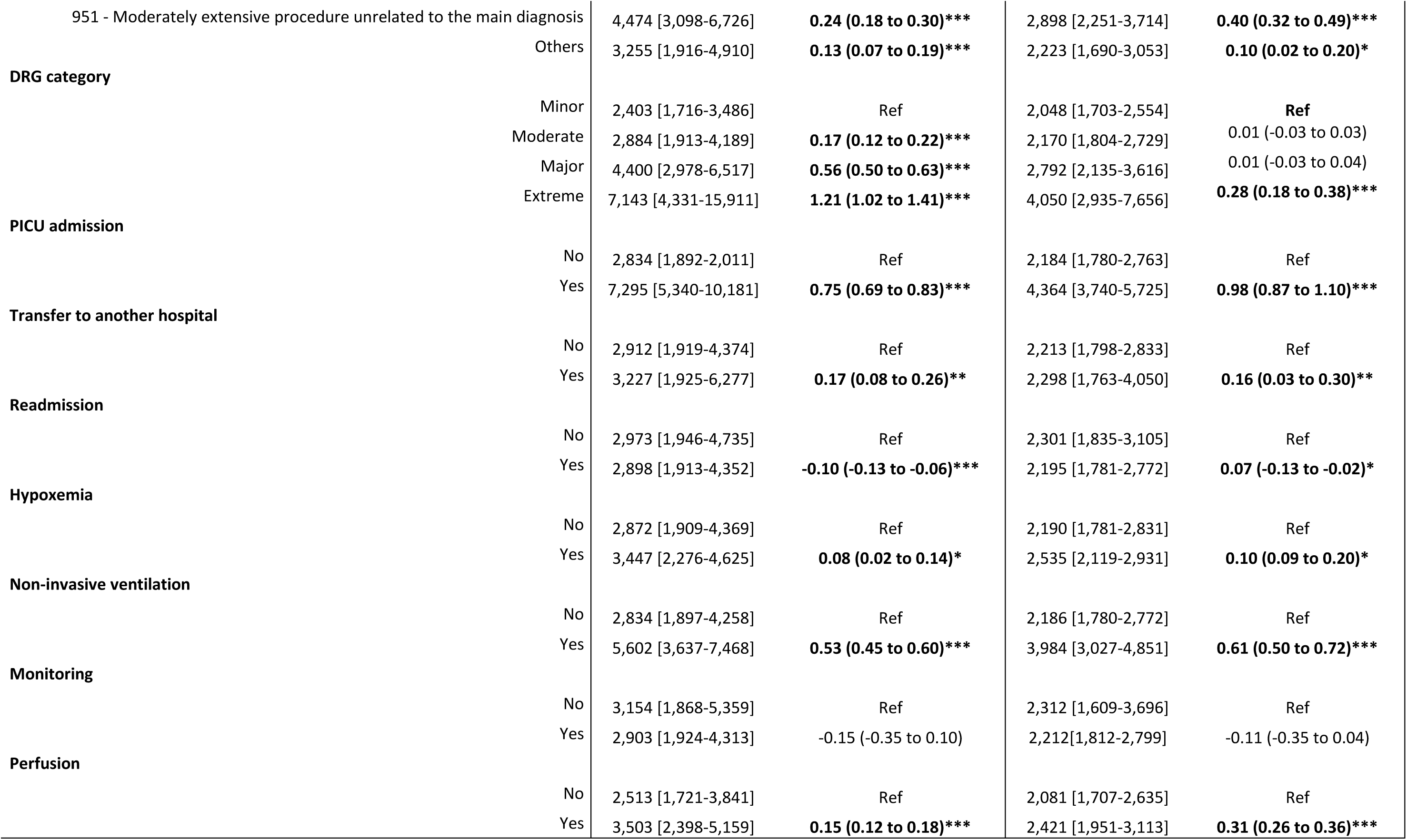
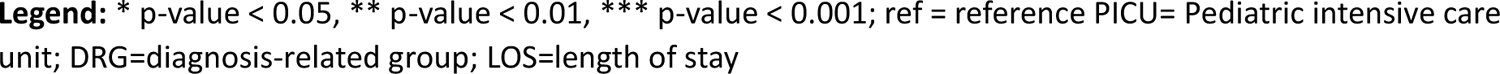
Factors associated with costs from the hospital and health insurance perspectives.

